# COVID-19 Infection in Children: Estimating Pediatric Morbidity and Mortality

**DOI:** 10.1101/2020.05.05.20091751

**Authors:** Michelle Barton, Kayur Mehta, Kriti Kumar, Jielin Lu, Nicole Le Saux, Margaret Sampson, Joan Robinson

## Abstract

**BACKGROUND:** Estimates of pediatric morbidity and mortality from COVID-19 are vital for planning optimal use of human and material resources throughout this pandemic.

**METHODS:** Government websites from countries with minimum 1000 cases in adults and children on April 13, 2020 were searched to find the number of cases confirmed in children, the age range, and the number leading to hospitalization, intensive care unit (ICU) admission or death. A systematic literature search was performed April 13, 2020 to find additional data from cases series.

**RESULTS:** Data on pediatric cases were available from government websites for 23 of the 70 countries with minimum 1000 cases by April 13, 2020. Of 424 978 cases in these 23 countries, 8113 (1.9%) occurred in children. Nine publications provided data from 4251 cases in 4 additional countries. Combining data from the websites and the publications, 330 of 2361 cases required admission (14%). The ICU admission rate was 2.2 *%* of confirmed cases (44 of 2031) and 7.2% of admitted children (23 of 318). Death was reported for 15 cases.

**CONCLUSION:** Children accounted for 1.9% of confirmed cases. The true incidence of pediatric infection and disease will only be known once testing is expanded to individuals with less severe or no symptoms. Admission rates vary from 0.3 to 10% of confirmed cases (presumably varying with the threshold for testing) with about 7% of admitted children requiring ICU care. Death is rare in middle and high income countries.

## Background

The 2019 Coronavirus disease (COVID-19) caused by the SARS-2 coronavirus came to world attention at the end of 2019 and was declared a pandemic by the WHO on March 11, 2020. The disease causes significant morbidity in adults where mortality rates are approximately 1 to 3%, although this figure is now approaching 8% in Italy.^1–3^ The case fatality rate appears to be heavily influenced by age and underlying medical conditions. Because almost all reported deaths are in adults, they have been the focus of preparedness for this pandemic. Describing the epidemiology of COVID-19 in children by providing preliminary data from countries with a significant number of cases will support predictions of the resources needed to adequately care for children and potentially allow redeployment of resources to other sectors if appropriate.

The objective of this paper was to estimate the current burden of COVID-19 infection in children, specifically the relative proportion of confirmed cases that occur in children, the need for hospital admission and the incidence of deaths in countries with significant COVID-19 activity, recognizing that this data will change over time.

## Methods

Two parallel searches were employed to address the study objectives.

### Web-based search

Countries with > 1000 confirmed COVID cases in adults and children as reported April 13 2020 on the WHO website were identified.^4^ The World Bank country income classification was used to assign an income status to each of these countries.^5^

Data from these countries were included in the study if a government website provided data on pediatric case numbers or deaths. Any available data on the age range of infected children, the number of pediatric hospitalizations, intensive care unit (ICU) admissions or deaths were recorded from included countries.

### Medical literature search

A literature search was conducted using the WHO database of COVID-19 literature, Embase, and MEDLINE on April 13, 2020. Records from all sources were consolidated using Mendeley, and then uploaded to COVIDENCE software for review (www.covidence.org). Search strategies are reported in Appendix 1. English and Chinese language publications were included.

The goal was to identify case series that could be used to supplement the data available from government websites. Studies originating from countries with more than 1000 cases on April 13 2020 were included if one or more of following could be derived for a group of minimum 20 infected children:

a. the age distribution of infected or admitted children
b. the number who required admission to hospital or to ICU
c. the number of deaths

Chinese language papers were translated by JL. Screening was performed by two authors. Discrepancies or disagreements were resolved by consensus.

Data were extracted on the total confirmed cases (defined as detection of COVID-19 from any site by any method), and the number and age range of pediatric cases, hospitalizations (or other measures of severity of illness) and deaths. As no standardized assessment of bias for such reports exists, none was performed.

### Data Analysis

Population-based data were compiled separately for each country and then combined for analysis as appropriate. Descriptive analysis was conducted and medians or means of continuous data were reported, where relevant. Sub-group analyses were done for different age ranges depending upon the availability of data.

## Results

### Web-based search

There were 70 countries with COVID-19 case counts exceeding 1000 on April 13 2020. They were of high (n=40), middle-high (n=22) and low-middle (n=8) income status. (Table 1). Pediatric data were available for 23 of the 70 countries from government websites (Table 2).

**Table 1.** Countries reporting more than 1000 adult and pediatric COVID cases on the World Health Organization website on April 13, 2020

|  | **Country** | **Income Status** | **Total Confirmed Cases** | **Total Deaths N (%)** | **Pediatric Data on Government Website** |
| --- | --- | --- | --- | --- | --- |
| 1 | United States of America | High | 524514 | 20444 (3.9) | yes |
| 2 | Spain | High | 166019 | 16972 (10.2) | yes |
| 3 | Italy | High | 156363 | 19901 (12.7) |  |
| 4 | Germany | High | 123016 | 2799 (2.3) | yes |
| 5 | France | High | 94382 | 14374 (15.2) |  |
| 6 | The United Kingdom | High | 84283 | 10612 (12.6) |  |
| 7 | China | Upper Middle | 83597 | 3351 (4.0) |  |
| 8 | Islamic Republic of Iran | Upper Middle | 71686 | 4474 (6.2) |  |
| 9 | Turkey | Upper Middle | 56956 | 1198 (2.1) |  |
| 10 | Belgium | High | 29647 | 3600 (12.1) |  |
| 11 | Netherlands | High | 25587 | 2737 (10.7) | yes |
| 12 | Switzerland | High | 25220 | 858 (3.4) | yes |
| 13 | Canada | High | 23702 | 674 (2.8) | yes |
| 14 | Brazil | Upper Middle | 20727 | 1124 (5.4) |  |
| 15 | Russian Federation | Upper Middle | 18328 | 148 (0.8) |  |
| 16 | Portugal | High | 16585 | 504 (3.0) |  |
| 17 | Austria | High | 13937 | 350 (2.5) | yes |
| 18 | Israel | High | 10878 | 103 (0.9) |  |
| 19 | Republic of Korea | High | 10537 | 217 (2.1) | yes |
| 20 | Sweden | High | 10483 | 899 (8.6) | yes |
| 21 | Ireland | High | 9655 | 334 (3.5) | yes |
| 22 | India | Lower Middle | 9152 | 308 (3.4) |  |
| 23 | Ecuador | Upper Middle | 7466 | 333 (4.5) | yes |
| 24 | Japan | High | 7255 | 102 (1.4) |  |
| 25 | Chile | High | 7213 | 80 (1.1) |  |
| 26 | Peru | Upper Middle | 6848 | 181 (2.6) |  |
| 27 | Poland | High | 6674 | 232 (3.5) |  |
| 28 | Norway | High | 6415 | 103 (1.6) | yes |
| 29 | Australia | High | 6322 | 61 (1.0) | yes |
| 30 | Romania | Upper Middle | 6300 | 306 (4.9) |  |
| 31 | Denmark | High | 6174 | 273 (4.4) | yes |
| 32 | Czechia | High | 5991 | 138 (2.3) |  |
| 33 | Pakistan | Lower Middle | 5374 | 93 (1.7) | yes |
| 34 | Malaysia | Upper Middle | 4683 | 76 (1.6) |  |
| 35 | Philippines | Lower Middle | 4648 | 297 (6.4) |  |
| 36 | Saudi Arabia | High | 4462 | 59 (1.3) |  |
| 37 | Indonesia | Lower Middle | 4241 | 373 (8.8) |  |
| 38 | Mexico | Upper Middle | 4219 | 273 (6.5) |  |
| 39 | United Arab Emirates | High | 4123 | 22 (0.5) |  |
| 40 | Serbia | Upper Middle | 3630 | 80 (2.2) |  |
| 41 | Luxembourg | High | 3281 | 66 (2.0) |  |
| 42 | Panama | High | 3234 | 79 (2.4) |  |
| 43 | Ukraine | Lower Middle | 3102 | 93 (3.0) |  |
| 44 | Qatar | High | 2979 | 7 (0.2) |  |
| 45 | Finland | High | 2974 | 56 (1.9) | yes |
| 46 | Dominican Republic | Upper Middle | 2967 | 173 (5.8) | yes |
| 47 | Colombia | Upper Middle | 2709 | 100 (3.7) | yes |
| 48 | Thailand | Upper Middle | 2579 | 40 (1.6) |  |
| 49 | Belarus | Upper Middle | 2578 | 26 (1.0) |  |
| 50 | Singapore | High | 2532 | 8 (0.3) |  |
| 51 | South Africa | Upper Middle | 2173 | 25 (1.2) |  |
| 52 | Greece | High | 2114 | 98 (4.6) | yes |
| 53 | Egypt | Lower Middle | 2065 | 159 (7.7) |  |
| 54 | Argentina | Upper Middle | 1975 | 82 (4.2) | yes |
| 55 | Algeria | Upper Middle | 1914 | 293 (15.3) | yes |
| 56 | Iceland | High | 1701 | 8 (0.5) | yes |
| 57 | Republic of Moldova | Lower Middle | 1662 | 33 (2.0) | yes |
| 58 | Morocco | Lower Middle | 1661 | 118 (7.1) |  |
| 59 | Croatia | High | 1600 | 23 (1.4) |  |
| 60 | Hungary | High | 1458 | 109 (7.5) |  |
| 61 | Iraq | Upper Middle | 1352 | 76 (5.6) |  |
| 62 | Estonia | High | 1309 | 25 (1.9) | yes |
| 63 | Kuwait | High | 1234 | 1 (0.1) |  |
| 64 | Slovenia | High | 1205 | 53 (4.4) |  |
| 65 | Bahrain | High | 1136 | 6 (0.5) |  |
| 66 | Azerbaijan | Upper Middle | 1098 | 11 (1.0) |  |
| 67 | New Zealand | High | 1064 | 5 (0.5) | yes |
| 68 | Lithuania | High | 1062 | 24 (2.3) |  |
| 69 | Armenia | Upper Middle | 1039 | 14 (1.3) |  |
| 70 | Bosnia and Herzegovina | Upper Middle | 1007 | 38 (3.8) |  |

**Table 2:**
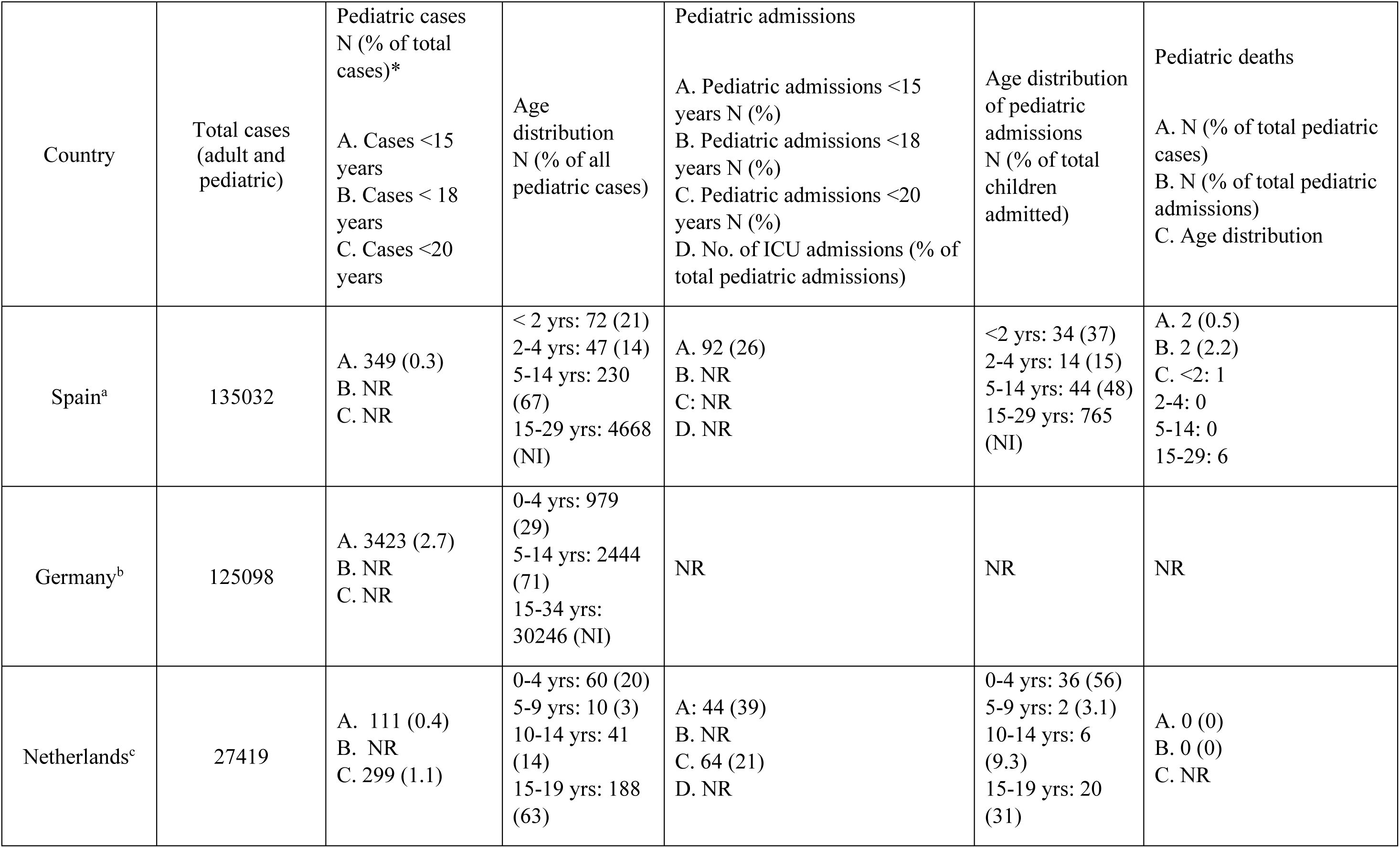
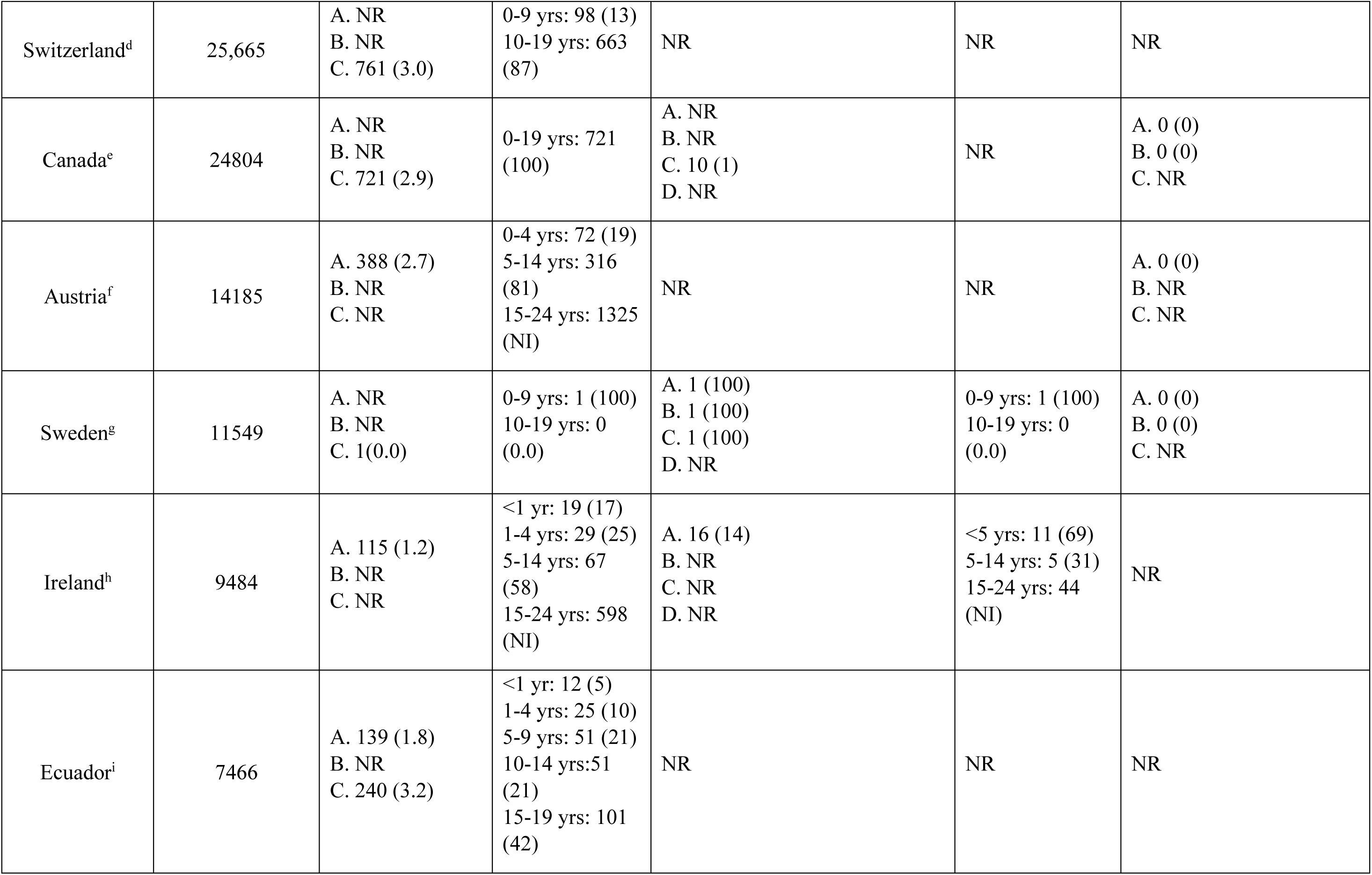
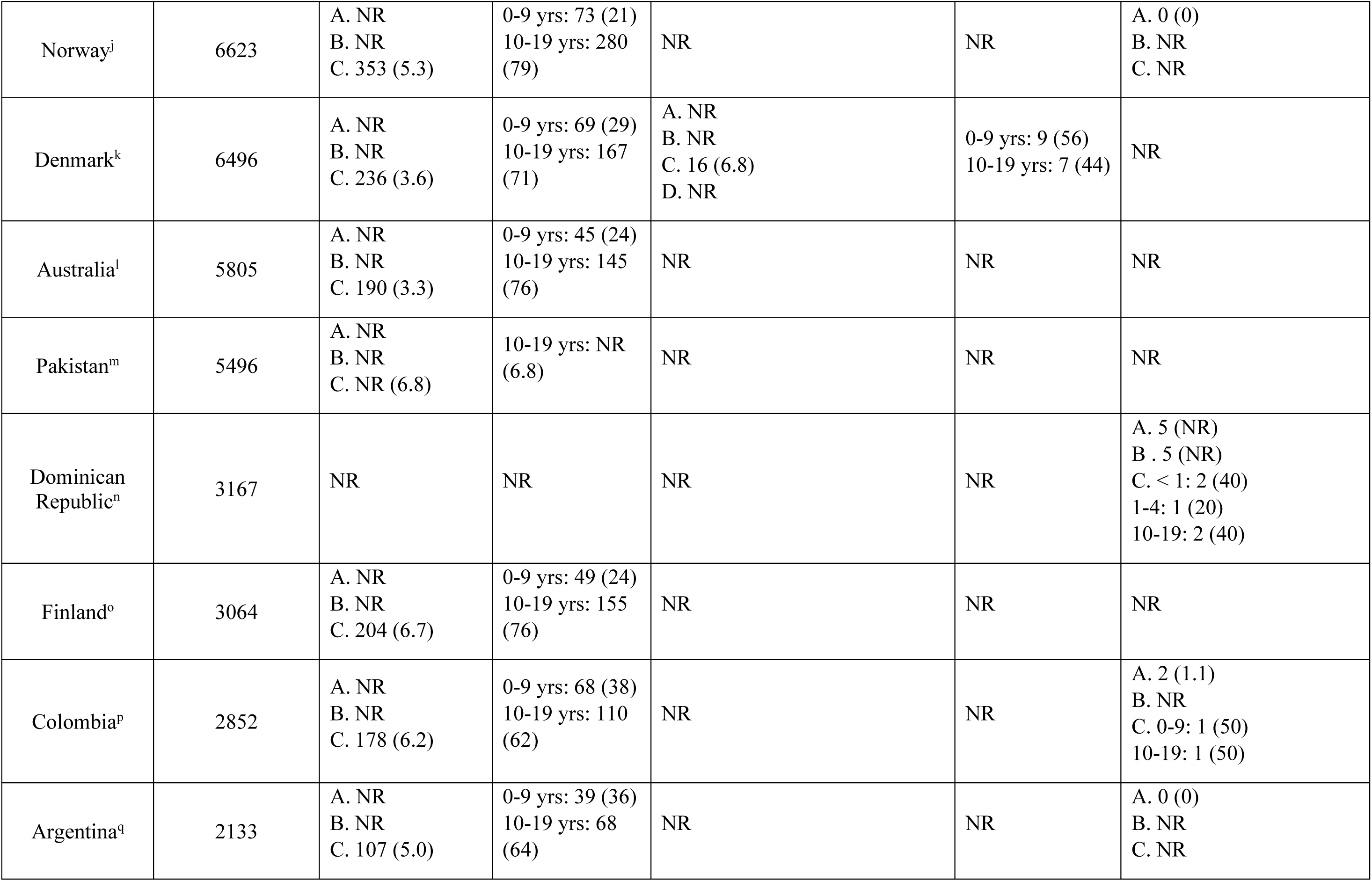
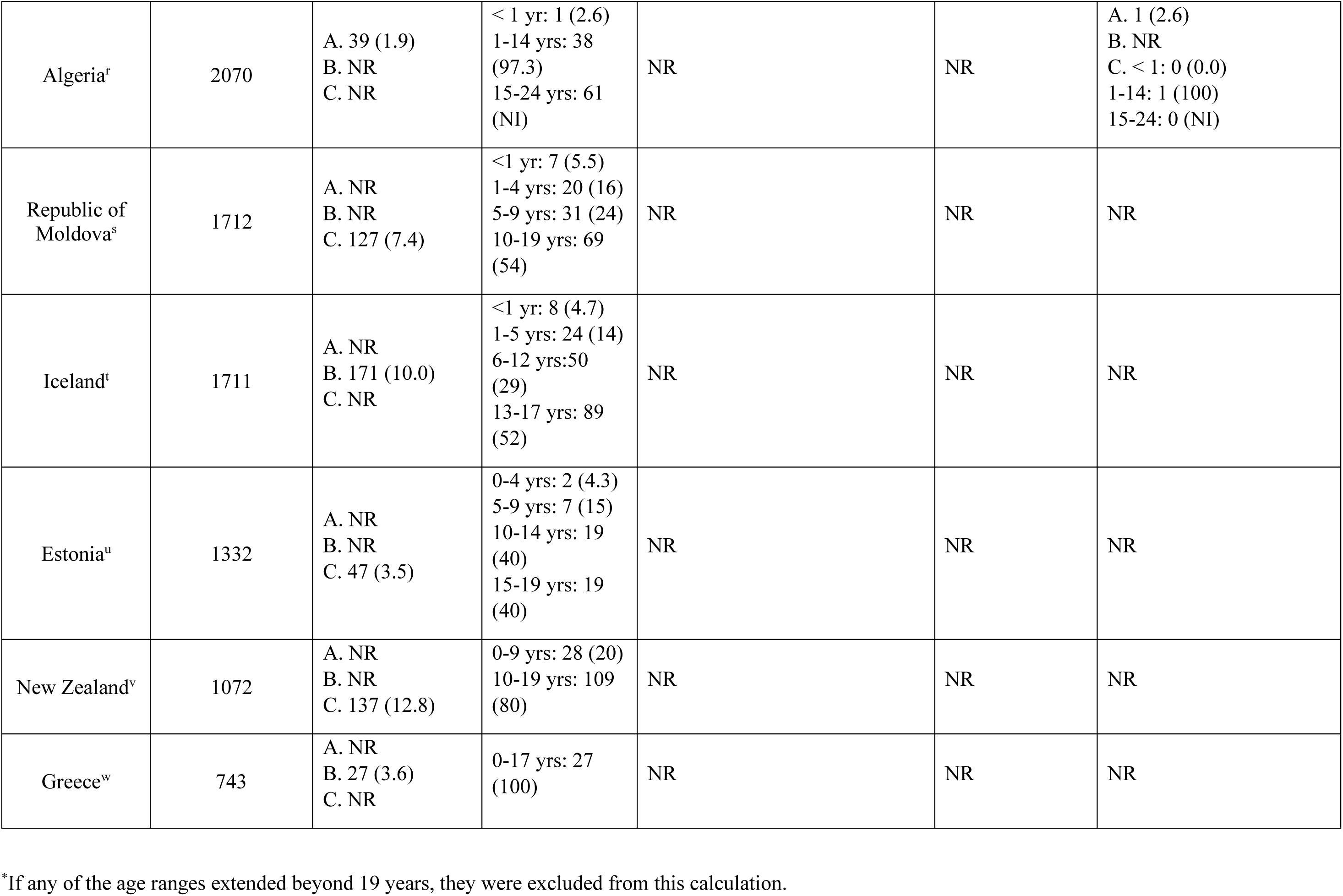
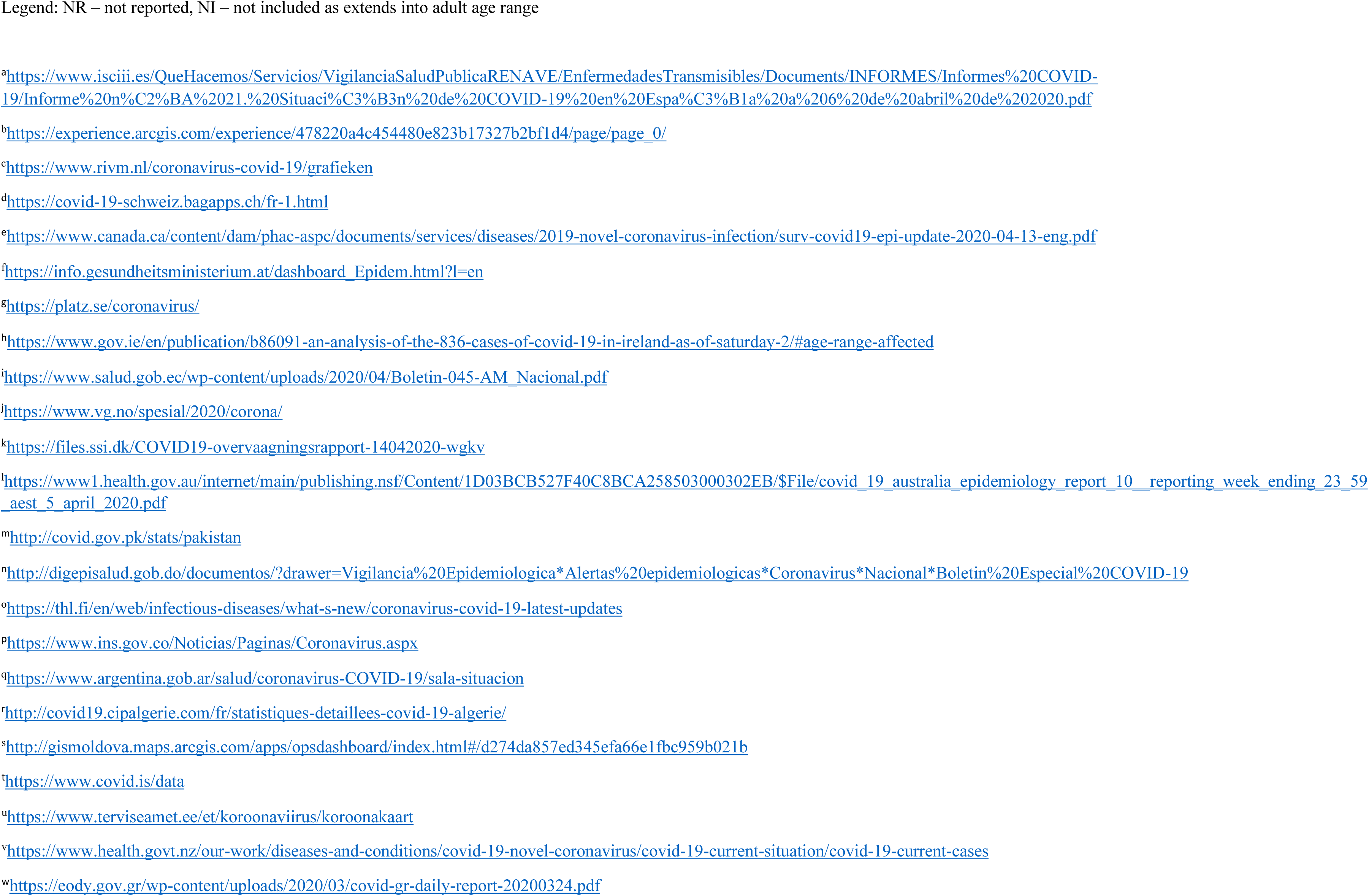
Burden of pediatric disease as obtained from government websites from countries reporting more than 1000 adult and pediatric COVID cases on the World Health Organization website on April 13, 2020

#### Relative proportion of pediatric cases

There were 424 978 COVID-19 infections reported for the 23 countries of which 8113 (1.9%) were pediatric (with the upper limit of the pediatric age range varying from 14 years to 19 years). Children accounted for a median of 3.5% (range 0.3-12.8%) of confirmed cases.

#### Age range and morbidity of infected children

There were 8 countries that reported the number of cases < 5 years of age, accounting for 1345 of 3262 cases < 15 years of age (41%) (Table 2). Admission occurred in 183 of 1606 children < 20 years of age in the 8 countries that reported this data (11%). The number of ICU admissions was not reported from any country. Death occurred in 5 of 5501 cases that occurred in countries where the number of pediatric deaths was stated on the website (0.09%). Another 5 deaths occurred in the Dominican Republic but the total number of infected children was not specified. ^6^

### Medical literature search

The search identified 700 unique titles from which 66 full text manuscripts were pulled (Figure 1) leading to 9 included case series with data on 4251 children from 4 countries (Table 3) ^7-18^ none of which were included in Table 2.

**Figure I.**
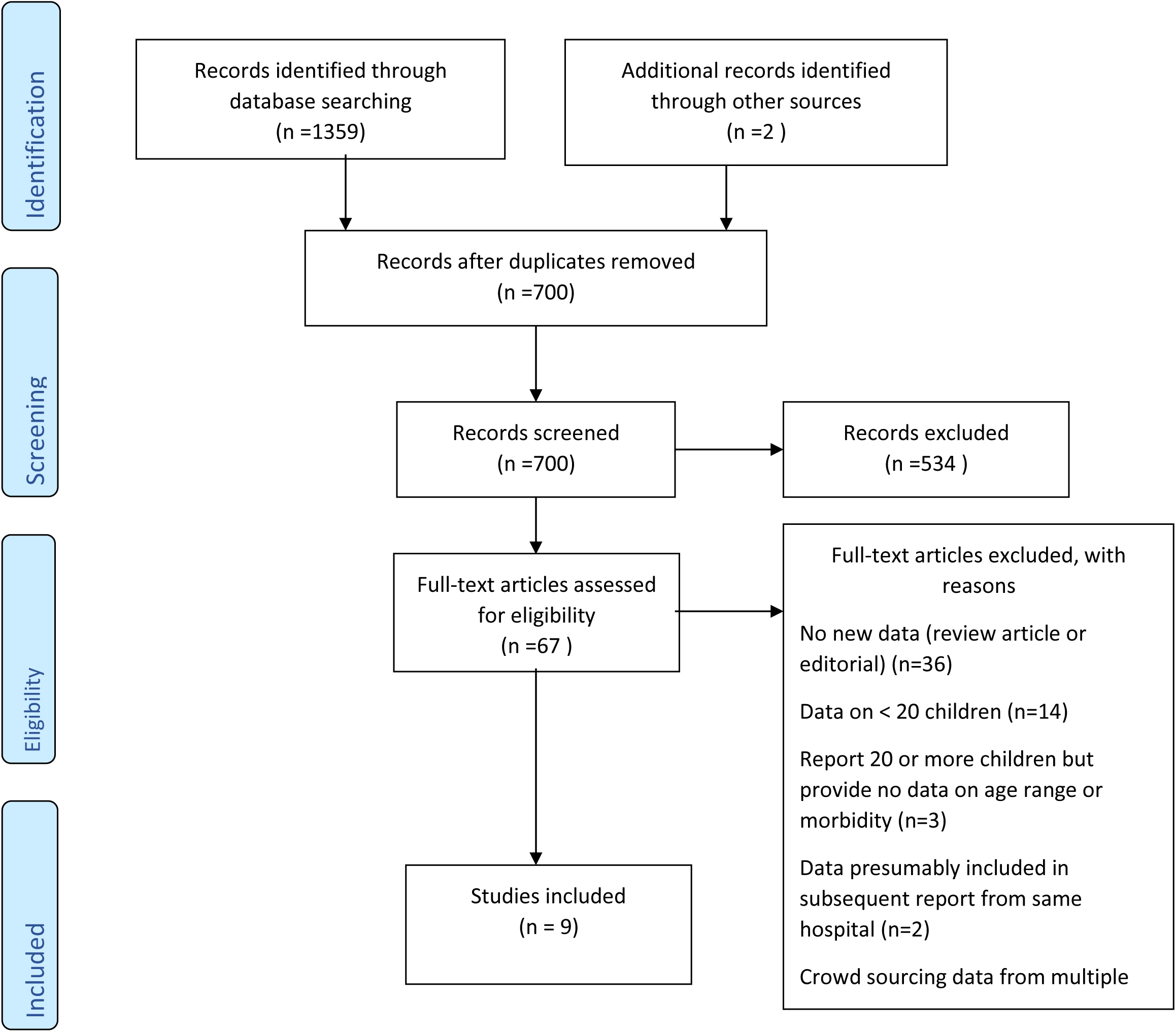
PRISMA diagram for systematic review for case series with minimum 20 children that provided data on age range or morbidity

**Table 3.**
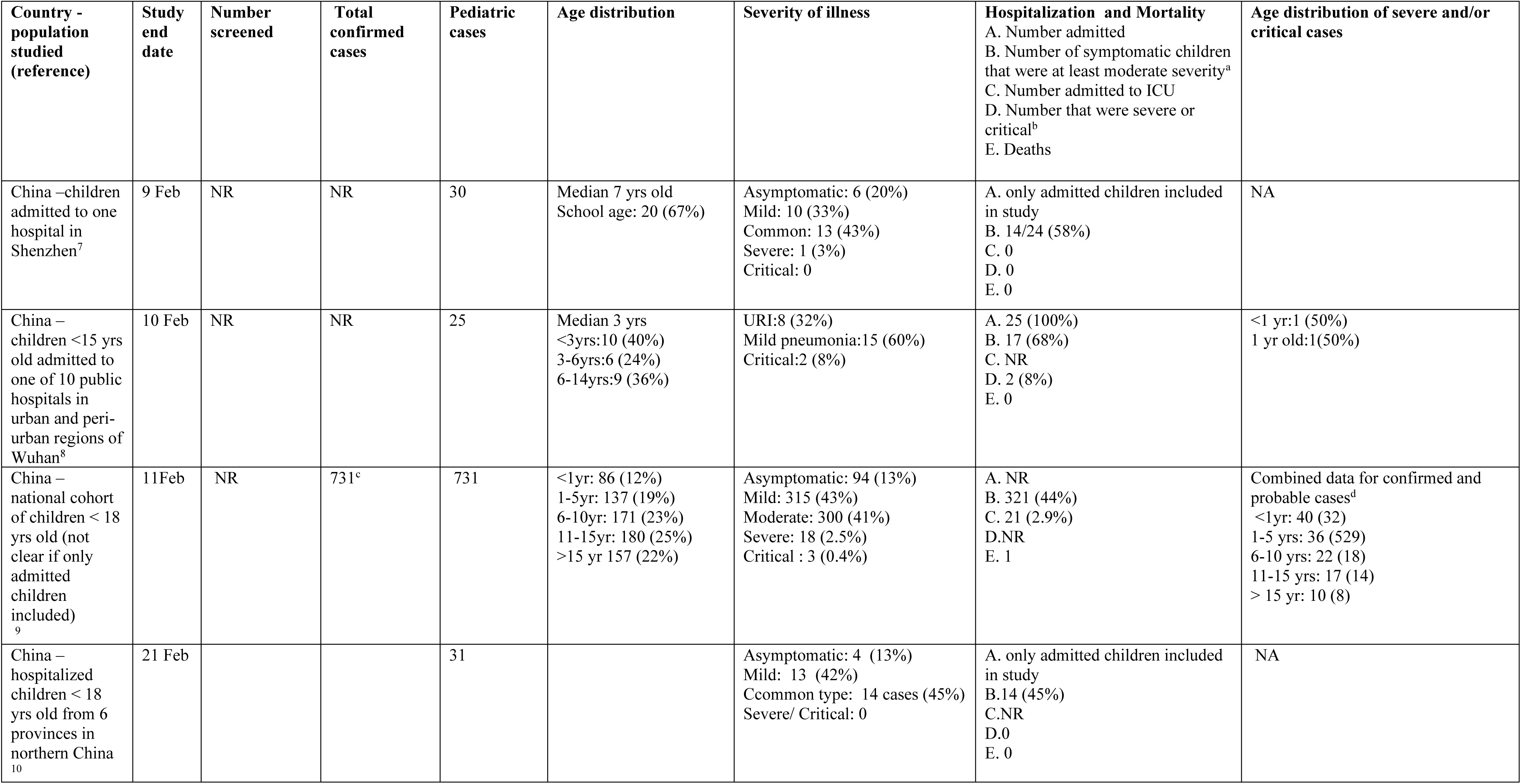
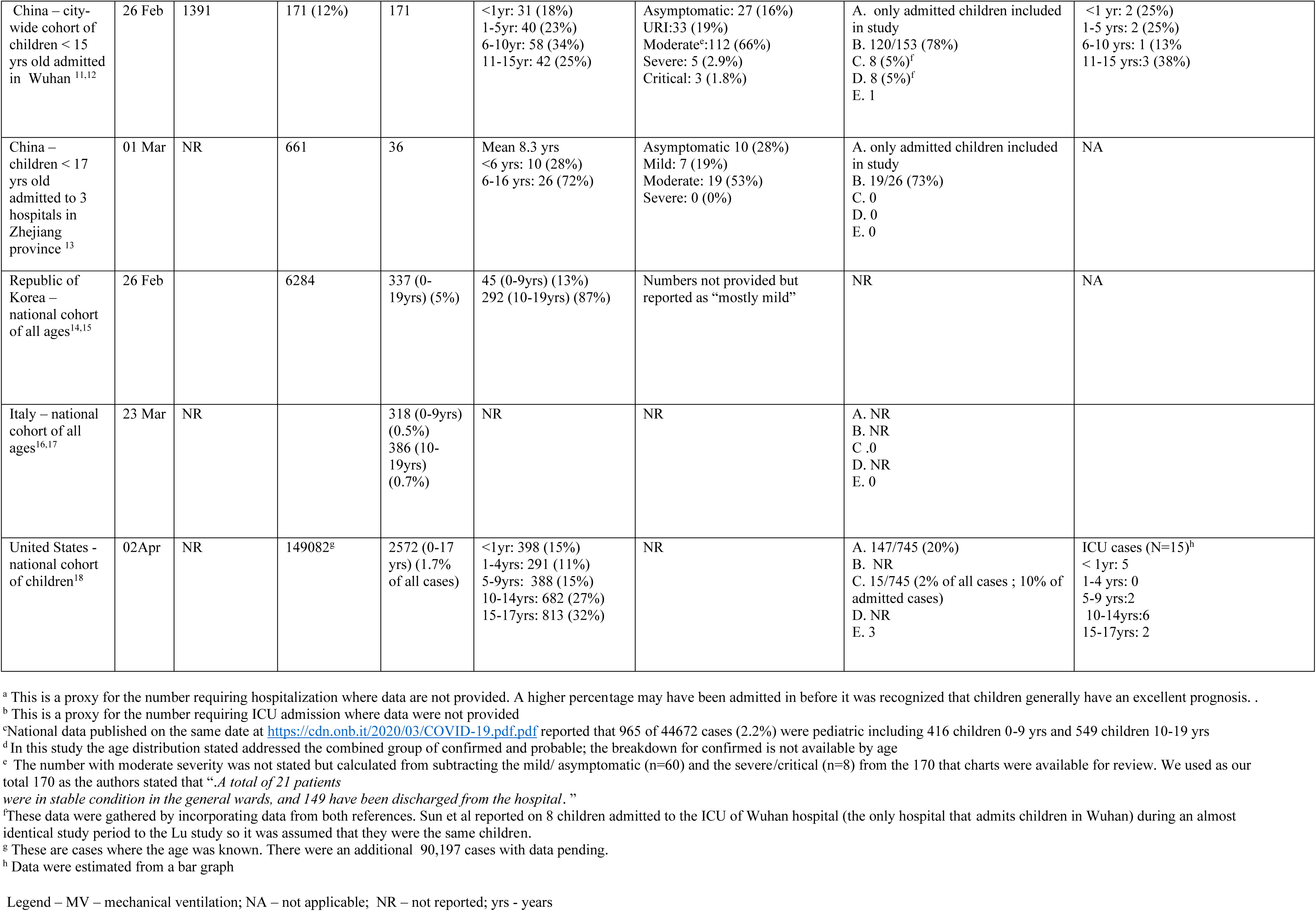
Details of pediatric COVID-19 case series with minimum 20 cases from review of the literature April 2 2020 (with US study updated April 6 2020)

#### Estimates of hospitalization and ICU admissions

The majority of infected children had mild disease. The hospitalization rate was 147 of 745 cases (20%) (Data were provided only for the US study). Moderate to severe disease which would presumably usually require hospitalization was reported in 44% to 78% of infected children for the other case series that included both inpatients and outpatients (Table 3). The ICU admission rate was 44 of 2031 confirmed cases (2.2%) (6 studies) and 23 of 318 admitted children (7.2%) (2 studies) and ranged from 2% to 8%. Three of 8 children in the smaller study reporting need for ICU in admitted children (from China) required mechanical ventilation ^11,12^ while data are not provided in the larger US report.^18^ For the 4251 cases in Table 3, there were 5 deaths reported (0.12%) although death may have occurred after these preliminary reports. The contribution of COVID-19 infection to death is not reported for these cases (a 10 month old infant with intussusception and a 14 year old male in China ^9,11^ and 3 US cases with details pending).^18^

#### Correlation between age and severity of disease

Children < 1 year of age accounted for 40 of 125 children (32%) with severe or critical disease if one combined presumptive and confirmed cases (data are not provided for confirmed cases alone) in the largest study from China.^9^ They accounted for 31 of 171 admissions (18%) and 25 of 111 cases of pneumonia (23%) in a smaller study from China and 2 (25%) of 8 ICU admissions.^11,12^ In the US, 59 of 95 children <1 year of age (62%) were admitted with 5 (5% of all cases and 8% of hospitalized cases) requiring ICU admission. ^18^

#### Combining data from the websites and from the publications

Admission occurred in 330 of 2361 cases (14%) where data were provided. There were minimum 15 deaths (not all websites and studies provided this data) in approximately 12 364 cases (the total number of cases is missing for the Dominican Republic where 5 deaths occurred) (0.12%).

## Discussion

Compared to adults, young children typically have a higher incidence of symptomatic infection with respiratory viruses due to their high exposure rates and lack of pre-existing immunity.^19^ To date, the pattern of disease secondary to the novel SARS CoV-2 virus appears to be an exception to this rule. Children accounted for approximately 2% of confirmed cases in the current study, although they make up 18% to 22% of the population in Korea, China and the United States.^15,18,20^ Admission rates for infected children would be predicted to vary markedly depending upon the criteria for testing and for admission. When COVID-19 was first recognized, almost all infected children were admitted due to the severity of disease in adults. However, it has become apparent that most children can safely remain as outpatients. The preliminary admission rate was 20% in 745 cases in the US^18^ but they estimate that the true rate could be as low as 5.7% since data were missing for 1827 cases and hospitalization status is more likely to be known for more severe cases. ICU admission occurred in about 5% of pediatric hospitalizations in a hospital in Wuhan, but the rate would have been higher had children with mild disease not been admitted.^11,12^ The preliminary ICU admission rate in the US appears to be higher at about 10% of hospitalized children but could be lower as ICU status is more likely to be known for severe cases.^18^ However, this rate could also increase as some reported US cases will still be in hospital. The criteria for ICU admission vary by hospital and must become stricter when there is a shortage of ICU beds. The death rate appears to be 0.1% to 0.2% in cases reported to date.

Children < 1 year of age seemed to account for a disproportionate percentage of admissions which could be because of clinicians having a lower threshold for admission of younger children.

It seems most likely that a large percentage of infected children are asymptomatic or manifest mild symptoms. The prevalence and the contribution of asymptomatic or mild infection to disease transmission is an emerging area of research.^21-23^ Deaths from COVID-19 predominate in the elderly and are commonly associated with a hyper-inflammatory state.^24^ It is not clear why this harmful immune response is rare in children. Understanding the presentations of COVID-19 disease in children and how they differ from the presentations of more common childhood viral illnesses should lead to a better understanding of the observed profound variation in morbidity and mortality between adults and children.^25^

The main limitation of this study is that pediatric data are yet to be reported for most countries, data would not have been recently been updated on all websites, and data on hospitalizations were missing from most websites. Data from the publications will already be out of date. The number of reported cases is very much determined by testing algorithms which may favor testing of elderly adults over children. The age ranges that were analyzed were not uniform with the upper limit for the pediatric age range varying from 14 to 19 years. The data in the current study may under-estimate ICU admissions and deaths. Initial data from China in adults with pneumonia showed a lag of about 12 days between symptom onset and admission so these outcomes may yet occur for children in the current study.^26^ The impact of COVID-19 on children will undoubtedly differ in low-income countries; intensive care is often not available and the proportion of children in the population is much higher.^27^

In conclusion, children account for a small percentage of symptomatic COVID-19 infections and severe and critical cases are rare. Further studies are required to delineate the true spectrum of pediatric disease worldwide and, perhaps even more importantly, the potential for children to develop severe disease if reinfection with COVID-19 or a similar coronavirus occurs later in life.

## Data Availability

Extracted data and meta-data for all bibliographic records screened is available on request.

## Appendix 1 Search Strategies

### Medline and Embase, Ovid multifile search

1. ((Wuhan adj3 (corona or virus)) or (ncov or COV 2 or 2019-nCoV or 2019nCoV or novel corona* or covid-19 or corona virus 2 or coronavirus* 2 or corona* 2019 or SARS-COV-2)).mp. (3534)
2. (baby* or babies* or newborn* or infan* or neonat* or preschool* or pre-school* or child* or pediatr* or paediatr* or teen* or adolescen* or youth).mp. (8960913)
3. 1 and 2 (261)
4. remove duplicates from 3 (166)
5. limit 4 to yr=“2019-Current” (102)

### Google Scholar

Google Scholar was searched March 20, 2020. The COVID-19 search concept was adapted from search developed by Wichor Bramer and posted on Search blocks/Zoekblokken (https://blocks.bmi-online.nl/catalog/397, visited March 18, 2020) Records were harvested using Publish and Perish

(covid19|”covid 19”|2019ncov|”2019 ncov|cov|coronavirus”|“2019 novel|new coronavirus|cov”|“wuhancoronavirus|cov|ncov|outbreak”|”wuhan*coronavirus|cov|ncov“|coronavirus|cov|ncov*wuhan”) AND (child|infant|adolescent|pediatric)

### WHO COVID-19 database

The World Health Organization (WHO) COVID-19 record set posted March 19, 2020 was downloaded from https://www.who.int/emergencies/diseases/novel-coronavirus-2019/global-research-on-novel-coronavirus-2019-ncov. The records were imported into Reference Manager, where topical queries were performed:

> All Indexed Fields {pediatric*} OR {paediatr*} OR {child} OR {infan*} OR {neonat*} OR { {adolesc*} OR {youth} OR {teen*}
>
> OR
>
> All Non-Indexed Fields {pediatric*} OR {paediatr*} OR {child} OR {infan*} OR {neonat*} OR { {adolesc*} OR {youth} OR {teen*}

